# Prospective exploration of a prognostic biomarker of nivolumab for head and neck cancer patients (BIONEXT)

**DOI:** 10.1101/2023.09.05.23295051

**Authors:** Kuniaki Sato, Satoshi Toh, Taku Murakami, Takafumi Nakano, Takahiro Hongo, Mioko Matsuo, Kazuki Hashimoto, Masashi Sugasawa, Keisuke Yamasaki, Yushi Ueki, Torahiko Nakashima, Hideoki Uryu, Takeharu Ono, Hirohito Umeno, Tsutomu Ueda, Satoshi Kano, Kiyoaki Tsukahara, Akihito Watanabe, Ichiro Ota, Nobuya Monden, Shigemichi Iwae, Takashi Maruo, Yukinori Asada, Nobuhiro Hanai, Daisuke Sano, Hiroyuki Ozawa, Takahiro Asakage, Takahito Fukusumi, Muneyuki Masuda

## Abstract

**BACKGROUND:** Nivolumab paved a new way in the treatment of patients with recurrent or metastatic (RM) head and neck squamous cell carcinoma (RM-HNSCC). However, the limited rates of long-term survivors (< 20%) demand a robust prognostic biomarker. This nationwide multi-centric prospective study aimed to identify a plasma exosome (PEX) mRNA signature, which serves as a companion diagnostic of nivolumab and provides a biological clue to develop effective therapies for a majority of non-survivors.

**METHODS:** Pre-treatment plasmas (*N* = 104) of RM-HNSCC patients were subjected to comprehensive PEX mRNA analyses for prognostic marker discovery and validation. In parallel, paired treatment-naïve tumor and plasma samples (*N* = 20) were assayed to elucidate biological implications of the PEX mRNA signature.

**RESULTS:** A combination of 6 candidate PEX mRNAs plus neutrophil-to-lymphocyte ratio precisely distinguished non-survivors from >2-year survivors (2-year OS; 0% vs 57.7%; *P* = 0.000124) with a high hazard ratio of 2.878 (95% CI 1.639-5.055; *P* = 0.0002348). In paired samples, PEX *HLA-E* mRNA (a non-survivor-predicting marker) was positively corelated with overexpression of HLA-E protein (*P* = 0.0191) and the dense population of tumor-infiltrating NK cells (*P* = 0.024) in the corresponding tumor, suggesting the HLA-E-NKG2A immune checkpoint may inhibit the antitumor effect of PD-1blockade in patients with high PEX *HLA-E* mRNA.

**CONCLUSION:** The PEX mRNA signature could be useful as a companion diagnostic of nivolumab. The combination of an anti-NKG2A antibody (i.e., monalizumab) and nivolumab may serve as a treatment option for non-survivors predicted by a RT-qPCR-based pre-treatment measurement of PEX mRNAs.

**TRIAL REGISTRATION:** This study is registered to the UMIN Clinical Trial Registry: UMIN000037029.

**FUNDING:** This study is partly funded by JSPS KAKENHI (Grant number JP 21436707 to MM) and Sota memorial fund to MM. PEXmRNA analyses were conducted by Showa Denko America Materials. CReS Kyushu organized sample collection and transfer, and conducted clinical data management with funding provided by Ono and Bristol-Myers Squibb.

**Graphical abstract:** 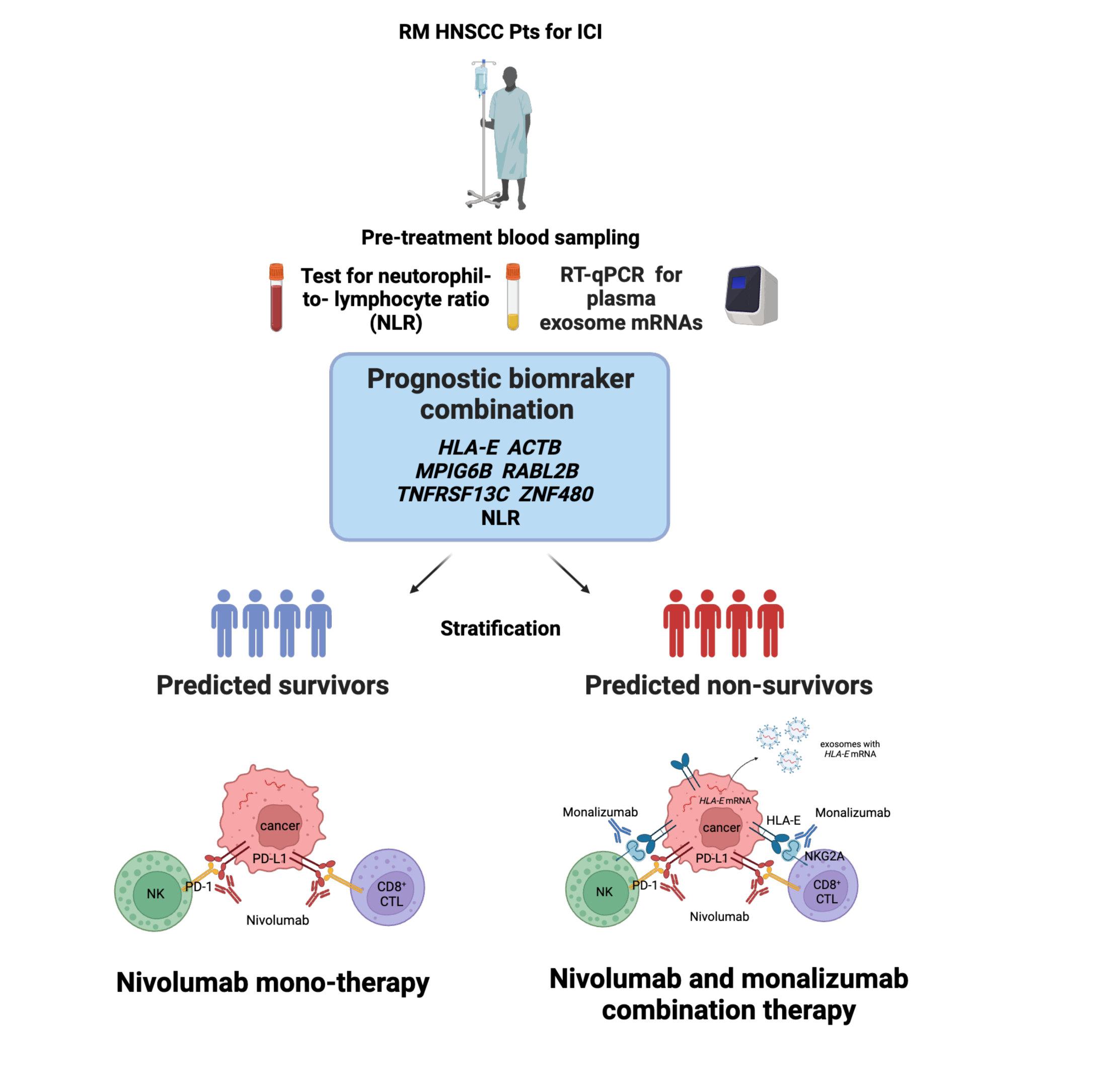

## Introduction

The emergence of immune checkpoint inhibitors (ICIs), especially those blocking programmed death-1 (PD-1), such as nivolumab or pembrolizumab, has had a substantial impact on the treatment of patients with recurrent or metastatic (RM) head and neck squamous cell carcinoma (HNSCC) (1). The CheckMate 141 study revealed that nivolumab treatments for selected patients achieved a long-term survival of >2 years for selected patients (2, 3), an unexpected achievement compared with conventional chemotherapeutic regimens. However, only 16.9% of patients experience this long-term survival (3); therefore, a reliable biomarker urgently needs to be established to address socioeconomic issues (4), and more importantly, an effective therapeutic strategy for a majority of non-survivors who don’t benefit from nivolumab administration needs to be developed.

The prognostic and predictive ICI biomarkers has been developed by the use of tissue sample-based methods including measurement of PD-L1 expression to determine the tumor proportion score (TPS) or combined positive score (CPS), tumor mutation burden, microsatellite instability, and interferon (IFN)-γ−related signatures (5–8). Overall, these indicators are utilized as a biomarker of pembrolizumab with limited clinical efficacy. In addition, these high cost, labor intensive, and time-consuming methods have insufficient accuracy for the response prediction of nivolumab and, more importantly, are not suitable to timely monitor the ever-changing tumor immune-microenvironment (TIME) of patients. It is necessary to establish a rapid and reliable biopsy-free prognostic biomarker (e.g., a biomarker that can be analyzed in blood) for nivolumab. In this context, exosome mRNA has attracted our attention. Exosomes are small-size (30-150 nm) extracellular vesicles secreted by a variety of cells, including cancer cells (9). Accumulating evidence indicates that exosomes function as cargos of biological information (i.e., proteins, lipids, DNAs, and RNAs), and significantly affect the milieus and physiological functions of the recipient cells in a context-dependent manner. Notably, exosome mRNAs are transcribed and function in the recipient cells (10). Exosomes mediate cross-talk between cancer cells and the extracellular matrix and normal cells (e.g., immune cells) and thus promote a specific tumor microenvironment that is advantageous for cancer cell proliferation, survival, migration, metastasis, and escape from immune surveillance (10). A recent milestone study demonstrated that exosomes secreted from *TP53*-mutated cancer cells can reprogram neurons into a cancer-promoting phenotype in HNSCC (11). The immune-suppressive effects of exosomes have also been confirmed in a series of HNSCC studies (12, 13). Thus, it is highly expected that the TIME of RM HNSCC, which regulates the response to nivolumab, can be assessed based on the plasma exosome (PEX) status. The isolation and quantification of exosome mRNA from patients is feasible in a couple of days. Therefore, we designed a multicentric prospective study to identify a PEX mRNA signature, which is measurable in clinical practice by reverse transcription-quantitative polymerase chain reaction (RT-qPCR). The main aim of this study is to establish a companion diagnostic for nivolumab that accurately predicts non-survivors and provides a clue for the development of a novel therapeutic strategy for non-survivors.

## Results

### Enrollment and clinical outcomes

The BIONEXT study is a multicentric prospective study involving 20 Head and Neck Surgery Departments in Japan and is composed of the two parts (Fig.1). The endpoint of part 1 of the study was the identification of a PEX mRNA signature that could segregate non-survivors from long-term (> 2-year) survivors. Part 1of the study enrolled 111 patients from July 7, 2019, to December 31, 2020, and the clinical data were collected and monitored until July 2022 by CReS Kyushu. Seven patients were excluded due to screening (*N* = 6) and sampling (*N* = 1) errors; therefore, the samples and clinical records of 104 patients were utilized for the biomarker assay and survival curve generation. Among them, 7 patients demonstrated a complete response (CR), 12 had a partial response (PR), 25 had stable disease (SD), 55 had progressive disease (PD), and 5 were not evaluated (NE) due to rapid tumor progression. The characteristics of the 104 patients are shown in Supplementary Table S1.

**Figure 1.**
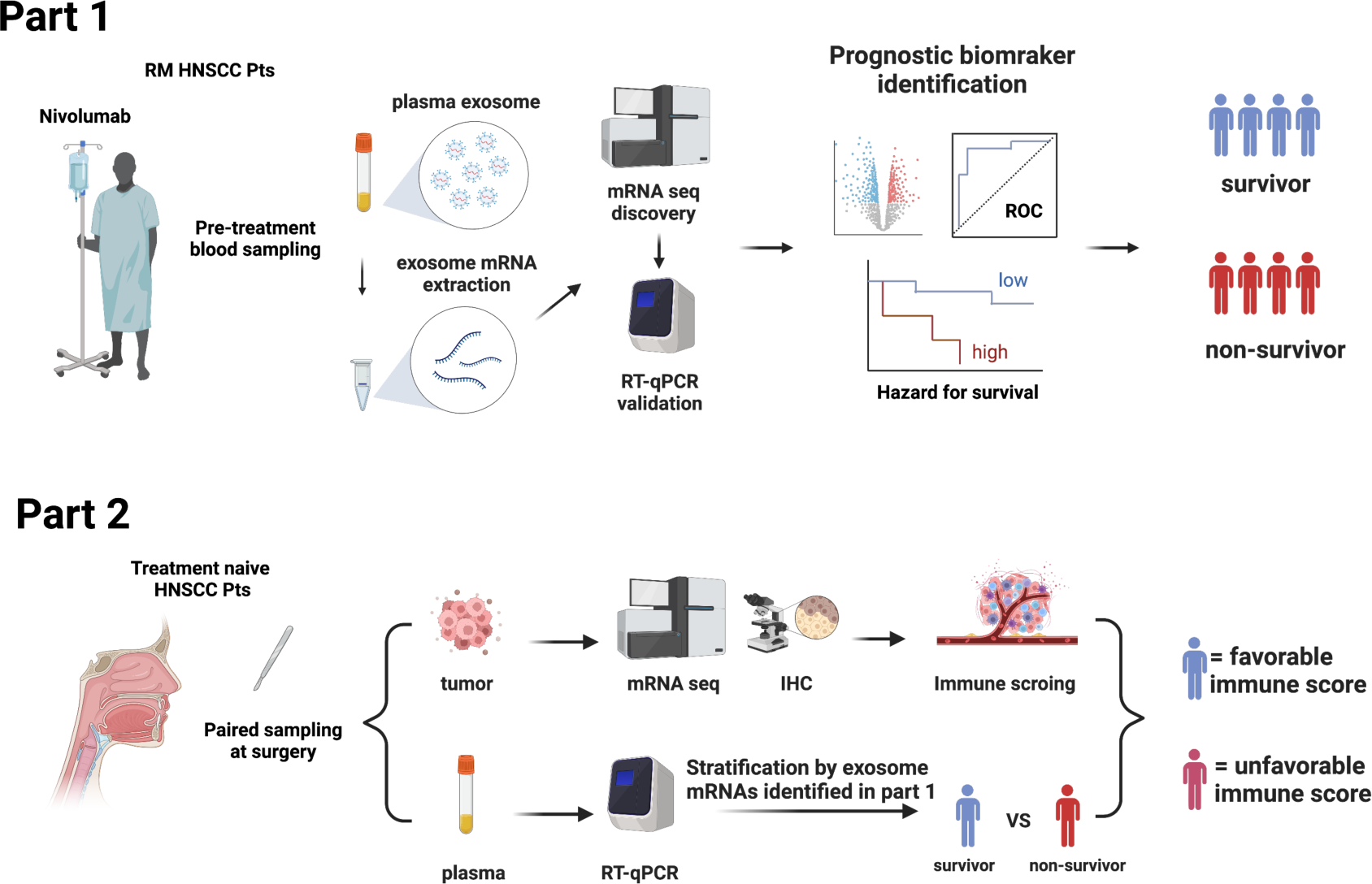
Study design.

### Candidate BOR-predicting PEX mRNA discovery

Based on the previous findings that the survival of patients treated by ICI could be stratified by best overall response (BOR)(14), we adopted a standard strategy to initially develop a BOR-predicting biomarker employing receiver operating characteristic (ROC) curve analyses and then to apply this biomarker to prognostic prediction by calculating cumulative survival rates and hazard ratios (HRs) between the marker-selected (i.e., high vs low) cohorts. In preparation for BOR-predicting biomarker exploration, we confirmed the accuracy of BOR for survival prediction in the current cohort (*N* =104). As shown in Figs. 2A and 2B, the overall survival (OS) rates of patients were well stratified in accordance with BOR; no patients were lost in the CR arm, while extremely poor prognosis was observed in patients with NE, who experienced rapid tumor progression before the first evaluation (Fig. 2A). Consequently, a substantial difference was found between the curves of responders (*N* = 19) and non-responders (*N* = 85) for 2-year OS (93.3% vs. 12.3%, Log-rank test *P* = 0.00000339; HR: 0.04; 95% confidence interval [CI]: 0.0055-0.293, *P* = 0.0015079) (Fig. 2B). However, not only responder (CR+PR), non-responder patients demonstrated long-survival; SD patients revealed a 48.7% of OS at 20 months and PD patients a 20.7% of OS at 2 years (Fig.2A), reflecting the fact that a certain portion of patients show durable responses to salvage chemotherapy beyond PD following nivolumab (15). Given that our main goal is to establish an accurate non-survivor-predicting biomarker, these beyond-PD survivors pose a conundrum. This is because, when response-predicting (i.e., responder vs non-responder) biomarkers are applied to survival analyses in this setting, a responder-predicting biomarker with high predictive value (specificity: score low patients = responders) keeps its power as a survivor-predicting prognostic biomarker, whereas a non-responder predicting biomarker with high predictive value (sensitivity: score high patients = non-responders) loses its power as a non-survivor-predicting prognostic biomarker, mis-predicting these beyond-PD survivors as non-survivors. Keeping this critical point in mind, we proceeded to the identification of a BOR**-**predicting PEX mRNA biomarker employing representative patients (PR: 6; SD: 5; and PD: 6) through comprehensive mRNA sequencing. We adopted a less restricted marker-selecting condition not confining the comparisons of groups in responders and non-responders and selected candidate BOR**-**predicting PEX mRNA, if they met one of the following criteria: 1) genes that were differentially expressed among the BOR categories (PR vs PD, PR vs SD/PD, and PR/SD vs PD) (*P* < 0.05), 2) genes with |log(fold change)| > 1.5, 3) genes with high area under the curve (AUC) values (> 0.7) in the ROC analyses for detection of PR vs PD, PR/SD vs PD (AUC1) and PR vs SD/PD (AUC2), or 4) genes identified as potential biomarkers in previous ICI studies (6, 8, 16, 17) or with high |log(fold change)| values in the present study. With these less broad criteria, the top 20 genes, *TAF4B, TESK2, MFSD8, RABL2B, ZNF480, FAM76A, TGIF1, TNFRSF13C, CTSW, LOC283788, SLC25A13, HLA-DQA1, COL10A1, MPIG6B, RPL23AP7, MSH2, CD3D, TCF7, HLA-E, and HLA-DRA* were selected as candidate BOR-predicting biomarkers for further analyses (Fig. 2C, and Supplementary Table S2).

**Figure 2.**
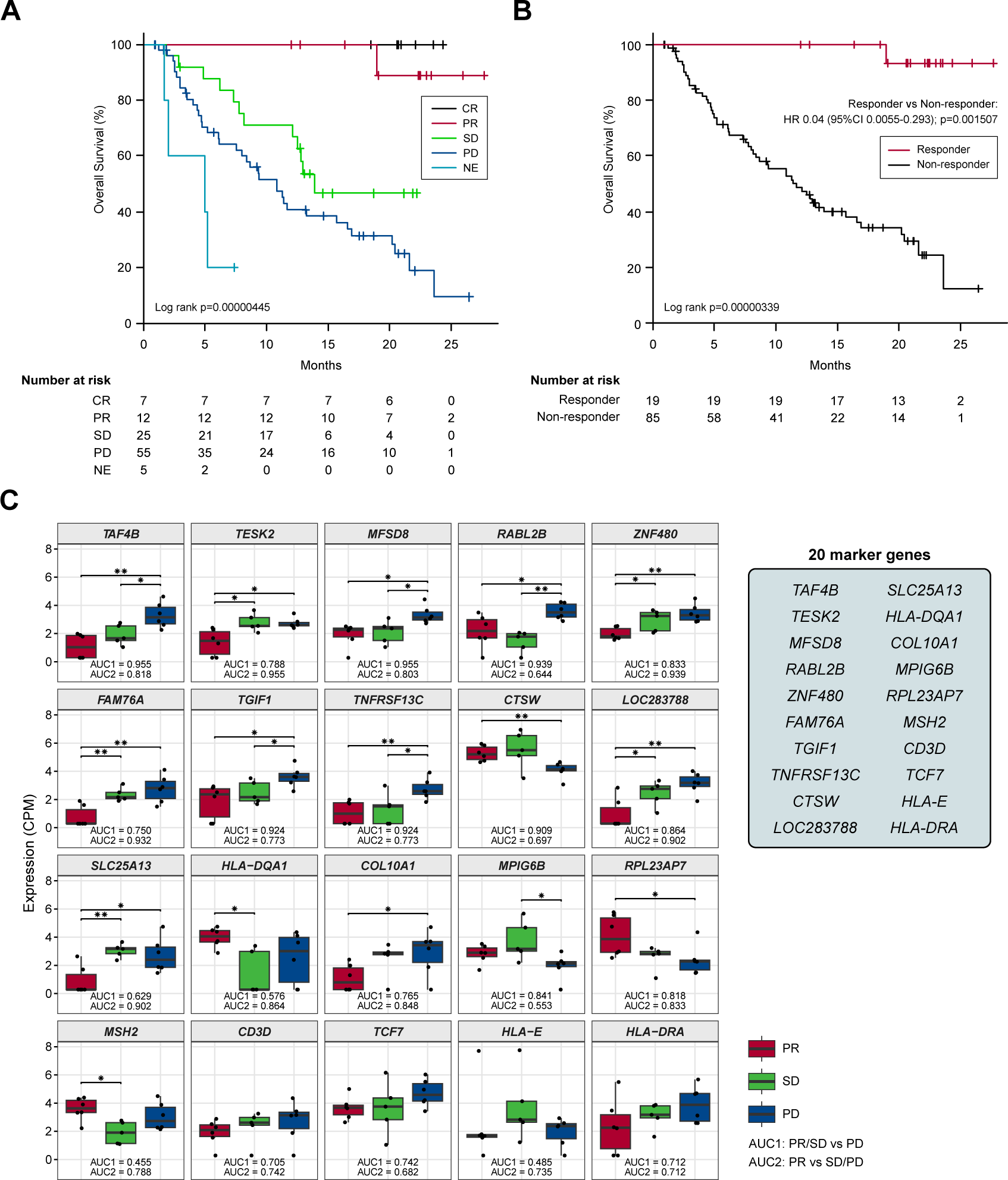
(A and B) Kaplan-Meier curves representing the overall survival of patients classified according to the best overall response. (C) Box plots representing the expression levels of 20 candidate biomarker genes in patients stratified according to the best overall response. CR, complete response; PR, partial response; SD, stable disease; PD, progressive disease; NE, not evaluated. (*) *P* <0.05; (**) *P* <0.005.

### Response-predicting PEX mRNA biomarker identification

Employing these candidate BOR-predicting PEX mRNAs, their powers for response prediction (i.e., responder vs non-responder) were investigated by RT-qPCR assays in the entire cohort (*N* = 104). To normalize the PEX mRNA data, two representative reference genes, *ACTB* and *GAPDH,* were added to assays. Interestingly, they demonstrated significantly (*ACTB, P* < 0.001; *GAPDH, P* < 0.05) higher expression (i.e., raw threshold cycle value) in the non-responder than in the responder. Assuming that these increases may reflect the vigorous total exosome production from aggressive cancer cells as confirmed in previous studies (10), we adopted *ACTB,* which had a greater difference, as one of the candidate biomarker PEX mRNAs, and used *GAPDH* as the reference gene. We then compared the expression levels of *GAPDH*-normalized 21 PEX mRNAs between responders and non-responders and their response-predicting powers were measured by the values of AUC in the ROC curve analyses and their optima thresholds were determined by the point nearest to the top-left corner on the ROC curve. The top 6 genes with the high AUCs, *HLA-E, ACTB, MPIG6B, RABL2B, TNFRSF13C,* and *ZNF480,* were selected as putative response-predicting biomarkers (Supplementary Table S3). PEX mRNAs that were increased in the non-responders (*HLA-E, ACTB, MPIG6B* and *TNFRSF13*) were considered as non-responder-predicting markers, while those that were increased in the responders (*RABL2B,* and *ZNF480*) were considered as responder-predicting markers (Fig. 3A). Of note, the *GAPDH*-normalized *ACTB* PEX mRNA remained significantly higher in non-responders. The AUC of these PEX mRNAs ranged from 0.593 to 0.729. For comparison, we calculated the AUC of the neutrophil-to-lymphocyte ratio (NLR), a proposed non-responder-predicting biomarker of ICI (18, 19), and found it was 0.591 (Supplementary Table S3 and Fig. 3A). The performance of individual PEX mRNAs was better than that of the NLR, but the values were not sufficiently high for clinical use. We then employed a simple algorithm to develop a better signature for response prediction by the combination of multiple PEX mRNA markers and the NLR. With the intent to generate non-responder-predicting combinations, we assigned 1 point if the expression of a non-responder-predicting gene or NLR exceeded the threshold or a responder-predicting gene fall below the threshold using the best threshold value (i.e., the point of the highest sensitivity and specificity) of each marker determined by the ROC curve analysis (Supplementary Table S3), and the points were averaged for various marker combinations. The score ranged between 0 and 1, and a score of 0 indicated that no marker in the combination predicted a non-responder, while a score of 1 indicated that all markers in the combination predicted a non-responder. To obtain the best combination of markers, we tested all the possible combinations of the top 6 markers and the NLR and identified the top 10 combinations with higher AUCs (ranging from 0.793 to 0.812) (Table 1 and Fig.3B). In the comparison of responders and non-responders, the scores of these combinations demonstrated more significant differences (*P* < 0.0005) (Fig.3B) than the mean expression of individual 6 PEX mRNAs and NLR, in which only *HLA-E*, *ACTB* (*P* < 0.05) and NLR (*P* < 0.005) demonstrated significant differences (Fig. 3A). Notably, all the top 10 combinations included *HLA-E*, which may suggest its importance for response prediction (Table 1).

**Figure 3.**
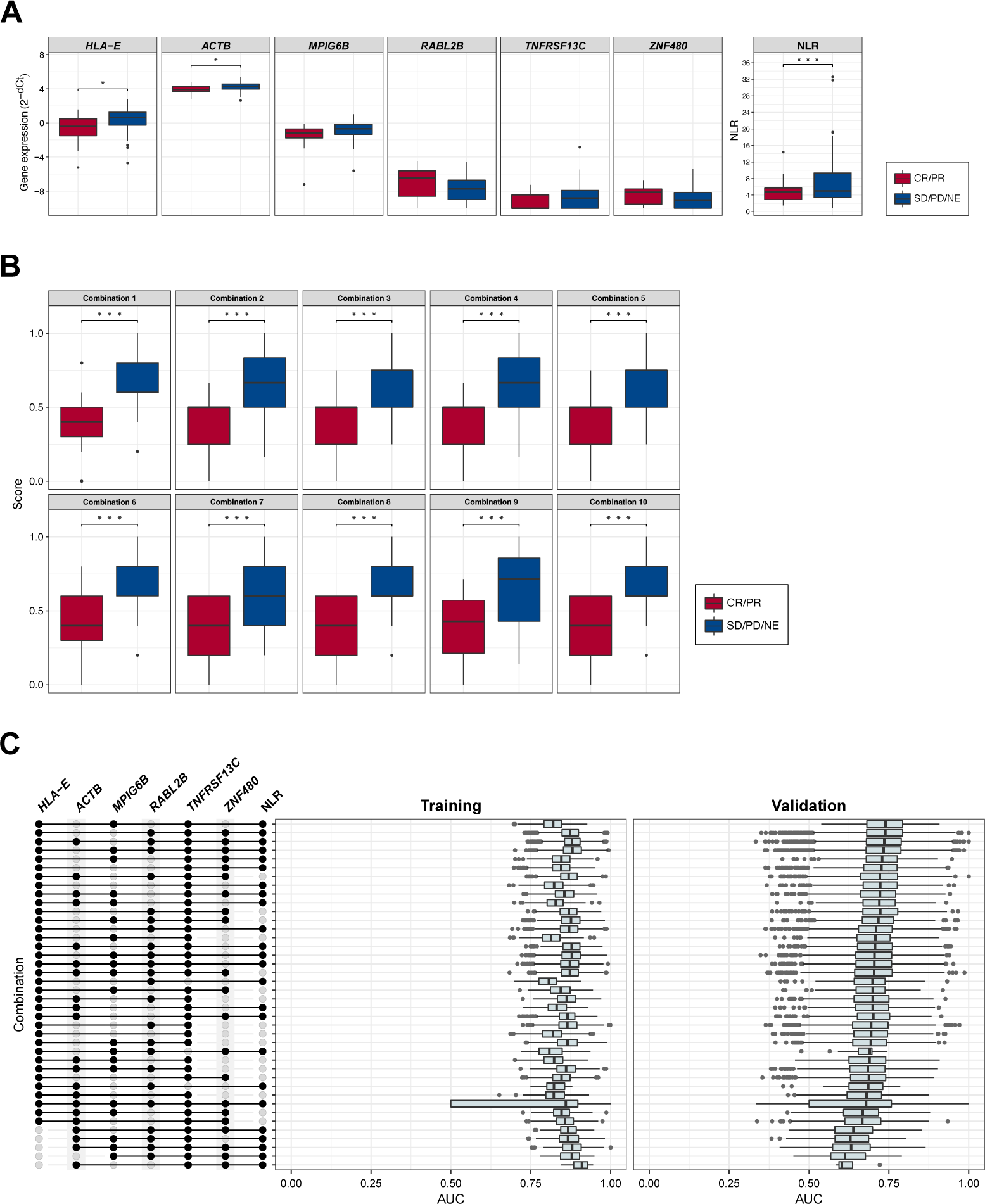
(A) Box plots comparing the expression levels of response-predicting biomarker genes and the neutrophil-to-lymphocyte ratios (NLR) between responders (CR/PR) and non-responders (SD/PD/NE). (B) Box plots comparing the scores of combinations calculated by biomarker genes and the NLR between responders (CR/PR) and non-responders (SD/PD/NE). (C) The area under curve (AUC) values for combinations of biomarker genes and the NLR (left panel, shown as an upset plot) in the training set (middle panel) and the validation set (right panel). (*) *P* <0.05; (**) *P* <0.005; (***) *P* <0.0005.

**Table 1.** Candidate response-predicting combinations (assessed in responder vs non-responder groups)

|  | AUC | threshold | sensitivity | specificity | ppv | npv | Markers |
| --- | --- | --- | --- | --- | --- | --- | --- |
| Comb 1 | 0.812 (0.716-0.907) | 0.5 | 0.765 | 0.737 | 0.929 | 0.412 | HLA-E RABL2B TNFRSF13C ZNF480 NLR |
| Comb 2 | 0.809 (0.722-0.896) | 0.583 | 0.635 | 0.895 | 0.964 | 0.354 | HLA-E ACTB RABL2B TNFRSF13C ZNF480 NLR |
| Comb 3 | 0.803 (0.714-0.893) | 0.625 | 0.635 | 0.895 | 0.964 | 0.354 | HLA-E RABL2B TNFRSF13C ZNF480 |
| Comb 4 | 0.801 (0.713-0.890) | 0.583 | 0.635 | 0.895 | 0.964 | 0.354 | HLA-E MPIG6B RABL2B TNFRSF13C ZNF480 NLR |
| Comb 5 | 0.796 (0.702-0.890) | 0.625 | 0.576 | 0.895 | 0.961 | 0.321 | HLA-E TNFRSF13C ZNF480 NLR |
| Comb 6 | 0.796 (0.702-0.890) | 0.5 | 0.765 | 0.632 | 0.903 | 0.375 | HLA-E ACTB RABL2B TNFRSF13C ZNF480 |
| Comb 7 | 0.795 (0.705-0.885) | 0.5 | 0.729 | 0.632 | 0.899 | 0.343 | HLA-E MPIG6B TNFRSF13C ZNF480 NLR |
| Comb 8 | 0.795 (0.703-0.887) | 0.5 | 0.753 | 0.579 | 0.889 | 0.344 | HLA-E ACTB TNFRSF13C ZNF480 NLR |
| Comb 9 | 0.794 (0.700-0.887) | 0.643 | 0.576 | 0.947 | 0.98 | 0.333 | HLA-E ACTB MPIG6B RABL2B TNFRSF13C ZNF480 NLR |
| Comb 10 | 0.793 (0.700-0.886) | 0.5 | 0.753 | 0.632 | 0.901 | 0.364 | HLA-E ACTB RABL2B TNFRSF13C NLR |

AUC, area under curve; ppv, positive predictive value; npv, negative predictive value

Before applying these response-predicting combinations to survival analyses, we tested the validity of our simple algorism for the generation of combination markers, comparing their AUC values to those obtained by a simulative sparse logistic regression algorithm (20). We selected samples to generate the same sample size (80%) as the study cohort by bootstrapping, trained the formulas using the selected samples (training), and validated the formulas in the remaining samples (20%) that were not used in the training (validation). The best marker combinations in each set, with high AUC values in the training and validation analyses, were selected after repeating this process for 3 sets at least 5,000 times each (Fig. 3C). High AUC values (0.819-0.923) were obtained in the training set, but only moderate AUCs (0.603-0.7418) were demonstrated in the validation set (Fig. 3C). This decreases of AUCs in the validation cohort indicated that our sample size was not sufficient to generate response-predicting gene combinations on a simulative platform. On the other hand, the AUC values obtained in our simple algorithm demonstrated well competitive values to those obtained in the training set. Considering its simplicity for the development of a companion diagnostic, the usefulness of this method was indicated. In addition, it was of note that *HLA-E* was selected at least 35 times out of the top 40 formulas, which corroborates the results of abovementioned combination approach.

### Prognostic biomarker identification

In our final assay, we investigated whether these non-responder-predicting combinations can serve as prognostic biomarkers for the prediction of non-survivors. Kaplan-Meier curves of patients were generated according to the thresholds of combinations 1-10 (Table 1). In combination 1, 2, 4, 7, 9, and 10, patients with high non-responder scores (above the threshold) demonstrated significantly (*P* = 0.0002348-0.0238) higher HRs (2.09-2.878) (Fig. 4A). Strikingly, in the most promising (i.e., high HR) combinations (9 and 10), the OS of the patients with high non-responder scores demonstrated a sharp drop towards 0% at 2 years (Fig 4B), while that of patients with low non-responder scores demonstrated an approximately 60% 2-year OS and a tendency to plateau after 20 months. Considering the highest HR and the lowest *P* value, we determined to adopt the combination 9 as a prognostic biomarker of nivolumab. Through the comparison of the survival curve of non-responder (*N* = 85) in Fig 2B and that of combination 9 score high patients (*N* = 50) in Fig.4B, we hypothesized that the beyond-PD survivors might be mainly involved in the score low cohort, improving the performance of the score for non-survivor-prediction. To test this hypothesis, we disassembled the Kaplan-Meier curve of combination 9 (Fig. 4B) based on the distribution of patients divided in the 2 x 2 contingency table (response x combination 9 score) (Fig. 4C) and compared the results to the BOR survival curve (Fig.2B). According to the table, non-responders (*N* = 85) were divided into 49 true positive and 36 false positive patients by the combination 9 score, causing a moderate sensitivity of 0.576 (49/85), a high ppv of 0.98(49/50), and a low npv of (0.333) (18/54) (Fig.4C and Table 1). These results in general indicated an imbalanced performance as a response-predicting biomarker. However, when the survival curves of the score-low-non-responders (false positive) and the score-high-non-responders (true positive) were compared, > 2-year survivors (i.e., the beyond-PD durable responders) were exclusively observed in the score-low group (36.8%) not in the score-high group (0%) (Fig.4C). Although these differences were not statistically significant (Log rank *P* = 0.0819, HR 1.642; 95% confidence interval 0.9335-2.887; *P* = 0.0852), this result supported our hypothesis. On the other hand, the responders (*N* = 19) were accurately grouped to the true negative score-low cases (*N* =18) showing a high specificity of 0.947 (18/19) and the survival curve of these score-low responders was identical with the responder curve segregated by BOR (Fig.4C and Fig.2B). The validity of our marker combination was confirmed by this improved specificity compared to those shown by the individual markers (0.526-0.684) (Table S3). In addition, the highest specificity of combination 9 (Table 1) suggested the superiority of this combination to other combinations. Taken together, the score-high group was composed of 49/50 (98%) non-responder-non-survivors and thereby the survival curve of the score-high led to the high prediction value of non-survivors (2-year OS: 0%), more accurately detecting the non-survivors than the survival curve of non-responders determined by BOR (2-year OS: 12.3%) (Fig.2B). Whereas, the score-low group included 18 responders with a 93 % of 2-year OS and 36 non-responders with a 36.8% of 2-year OS and the survival curve of the score-low patients resulted in the merged curve of these two populations resulting in a 57.7 % of 2-year OS (Fig.4B). Although being associated with unusual pattern of parameters, our biomarker demonstrated the high performance as a non-survivor-predicting biomarker, at least in part, by the ability to exclusively separate the beyond-PD durable responders into the score-low non-responders (Fig.4C). In conclusion, it was confirmed that a biomarker combination can segregate non-survivors from > 2-year survivors with relatively high accuracy and that the prediction of non-survivors is, to some extent, feasible with a single pretreatment RT-qPCR-based blood test.

**Figure 4.**
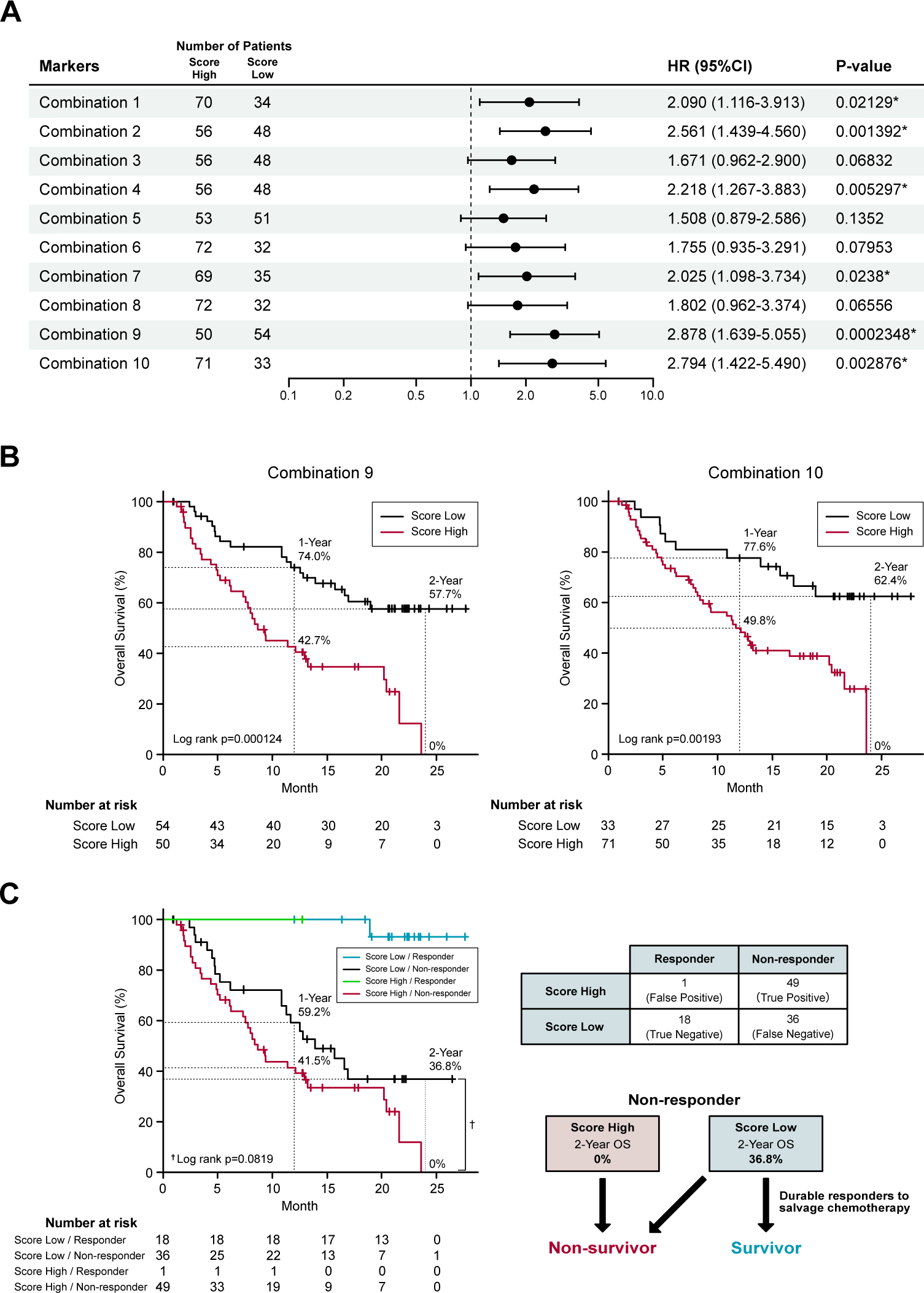
Survival prediction based on the identified biomarker combinations. (A) Forest plots representing the hazard ratios of the biomarker combinations. HR, hazard ratios; CI, confidence intervals. (B) Kaplan-Meier curves representing the overall survival of patients classified according to the score of biomarker combination (left panel, combination 9; right panel, combination 10). (C) Kaplan-Meier curves (left panel) representing the overall survival of patients classified according to the 2 x 2 contingency table (right panel).

### Correlation of prognostic biomarker combinations with the TIME

In part 1 of our study, we identified a prognostic biomarker combination (*HLA-E, ACTB, MPIG6B, RABL2B, TNFRSF13C, ZNF480* and NLR) that could precisely predict non-survivors treated with nivolumab. We then proceeded to part 2 of the study to confirm that the combination of 6 PEX mRNAs and NLR indeed reflect the TIME and, more importantly, to find a biological clue for the development of novel strategies for non-survivors (Fig. 1). For part 2, 20 paired blood, plasma and tumor samples were collected from the treatment-naïve HNSCC patients who underwent radical surgery at National Kyusyu Cancer Center. Blood samples were used for the measurement of NLR. Plasma samples were subjected to PEX mRNA assay and tumor samples were to RNA-seq and IHC. Sufficient tissue amounts for RNA-seq were not obtained for 3 frozen tumor samples; thus, 17 tumor samples were subjected to the mRNA analyses, 20 tumor samples were subjected to the IHC analysis, and 20 plasma samples for PEX mRNA assay. We first measured the expression levels of *GADPH*-normalized 6 PEX mRNAs by RT-qPCR and the levels of mRNAs in the corresponding tumors by RNA-seq to examine their correlations. Consistent with the previous finding that only specific genes demonstrated significant correlations (21), PEX *HLA-E* mRNA showed a near-significant (*P* = 0.052) correlation with tumor *HLA-E* mRNA among the 6 genes (Fig. 5A). In view of this positive tendency, we compared the expression levels of PEX *HLA-E* mRNA and HLA-E protein in the tumors and found a significant association (*P* = 0.0191) (Fig. 5C). Collectively, the high PEX *HLA-E* mRNA expression appears to reflect the high *HLA-E* mRNA transcription and protein translation in the corresponding tumor. We then attempted to stratify the 20 patients into score high candidate non-survivors and score low candidate survivors based on the biomarker combination 9 established in part 1 of the study (Table 1) (Fig.1A). However, interestingly, all 20 patients were grouped as survivors, because in the blood and plasma samples obtained from the treatment-naïve patients the NLR and the mean PEX mRNA expression levels of 6 PEX mRNAs except for *TNFRSF13C* indicated favorable response patterns compared to the RM samples (Fig. 5B): i.e., lower in the non-responder genes (*HLA-E, ACTB,* and *MPIG6B*) and higher in the responder genes (*RABL2B* and *ZNF480*). This result is consistent with the fact that the TIME of treatment-naïve tumor is more tumor-eliminating compared to the exhausted TIME of RM tumor, warranting the efficacy of this biomarker as a monitor of the TIME. Given the prominent role of HLA-E repeatedly identified in the present study and its importance as a target of immunotherapy (i.e., therapies targeting the HLA-E-NKG2A immune checkpoint) (22, 23), we alternatively utilized the mean value of PEX *HLA-E* mRNA to stratify the 20 patients. We compared the status of immune parameters (PD-L1 CPS score, IFN-γ-related signature score, and CIBERSORT-derived infiltrating immune cell levels) (6, 7, 24) between PEX *HLA-E* mRNA high (*N* = 10) and low (*N* = 10) patients. The CPS (*P* = 0.6242) and IFN-γ-related signature (*P* = 0.1802) did not show significant correlations with the levels of PEX *HLA-E* mRNA. However, the number of activated natural killer (NK) cells determined by CIBERSORT were significantly (*P* = 0.024) abundant in the tumors of patients with high PEX *HLA-E* mRNA (Fig. 5D). It is known that HNSCC is the most immune-infiltrating cancer types across the solid tumors (25) and these tumor-infiltrating NK cells and CD8^+^ cytotoxic T lymphocytes (CTL) strongly express NKG2A and PD-L1(23). Considering the positive correlation of PEX *HLA-E* mRNA and HLA-E protein expression confirmed above, the effects of PD-1blockade by nivolumab may be canceled by HLA-E-NKG2A check point in patients with high PEX *HLA-E* mRNA, as illustrated in Fig.5D. Thus, the combination of clinically usable anti-NKG2A antibody (i.e., monalizumab) and nivolumab may be useful for the candidate non-survivors predicted by the pre-treatment biomarker combination indicating high PEX *HLA-E* mRNA.

**Figure 5.**
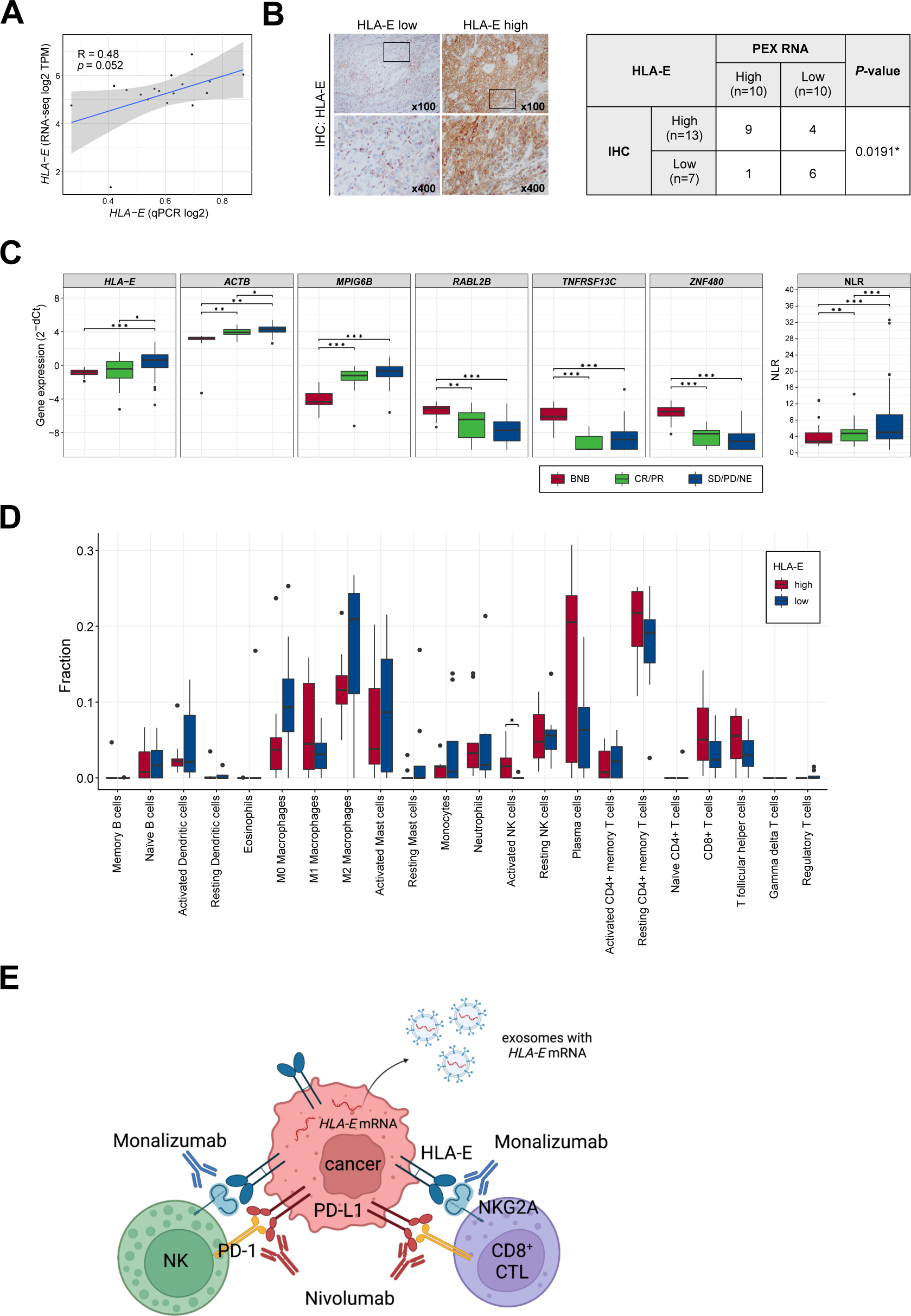
Correlation of PEX mRNA and the tumor immune microenvironment. (A) Correlation between the *HLA-E* expression levels detected by RNA-seq (vertical axis) and qPCR (horizontal axis) (*N* =17). R represents the Pearson correlation coefficient. (B) Representative images of immunohistochemistry staining for HLA-E in tumor tissues; HLA-E low (left) and HLA-E high (right). High-magnification images of the regions indicated by black boxes are shown. The table represents the numbers of cases and the correlation between HLA-E protein and *HLA-E* mRNA expression levels in PEXs (*N* =20). The P-value was calculated by Fisher’s exact test. (*) *P* <0.05; (**) *P* <0.005; (***) *P* <0.0005. (C) Box plots representing the expression levels of biomarker genes detected by RT-qPCR of exosomes extracted from peripheral blood and the NLR. BNB represents the cohort of part 2 study cohort (*N* =20). CR, complete response; PR, partial response; SD, stable disease; PD, progressive disease; NE, not evaluated. (D) Box plots representing the proportion of immune cells estimated by CIBERSORTx in the primary tumor tissues (*N* =17). Patients were classified according to the expression levels of PEX *HLA-E* mRNA (HLA-E high, *N* =9; HLA-E low, *N* =8). P-value was calculated by Mann-Whitney U-tests. (*) *P* <0.05; (**) *P* <0.005; (***) *P* <0.0005. (E) Schematic summarizing of our proposed mechanism by which the effect of nivolumab is canceled in the tumor of patient with high PEX HLA-E mRNA. The high PEX mRNA level reflects the vigorous HLA-E protein production in cancer cells, forming HLA-E/NKG2A checkpoint with NK and CD8+CTL cells. In this setting, administration of nivolumab alone is not effective. Addition of an anti-NKG2A antibody, monalizumab, is expected to restore the cytotoxic effects of NK and CTL cells circumvented by the dual immune checkpoints. (*) *P* <0.05; (**) *P* <0.005; (***) *P* <0.0005.

## Discussion

To the best of our knowledge, this is the first prospective study to demonstrate the feasibility of a single pretreatment RT-qPCR-based blood test for predicting the non-survivors with RM HNSCC treated with to nivolumab. In this study, we adopted a standard strategy to apply response-predicting biomarkers identified by the ROC curve to the survival analyses (26). However, as described in the manuscript, a substantial portion of non-responders demonstrates beyond-PD durable responses for salvage chemotherapy following nivolumab and therefore the development of an high-performance non-responder-predicting biomarker, (in this case with high sensitivity) may results in the poor performance as a prognostic biomarker mis-predicting these survivors as non-survivors before treatment: a phenomenon not associated with conventional chemotherapeutic agents (15). To address this challenging problem, we broadly curated BOR-predicting PEX mRNAs in the discovery cohort, expecting that this method may, to some extent, suppress sensitivity and thereby lead to the improved survival prediction. As shown in Fig.4C, this scheme contributed to the detection of the beyond-PD long-term survivors exclusively in the score-low non-responders. Although the precise mechanism of this segregation remains unknow, our results proved the efficacy of the response-predicting combination 9 as a prognostic biomarker when applied to the prediction of non-survivors, achieving our main goal.

Currently, an IFN-γ-related signature (the original 6 genes and an expanded 18-gene signature), which was established as a biomarker of pembrolizumab using the tissue-based NanoString platform, is often employed (5, 8) based on its relatively high AUC of 0.75 for response prediction in RM HNSCC (6). However, the power of this biomarker remains unclear when utilized as a prognostic biomarker. The present study revealed that our liquid biomarker combinations consisting of 6 PEX mRNAs and the NLR demonstrated similar AUC of 0.794 for response-prediction and as well showed high performance as a prognostic biomarker. Considering, the accuracy, speed and ease with which it can be assayed, the pretreatment measurement of NLR and PEX mRNA signature by RT-qPCR may be a novel companion strategy for nivolumab therapy in patients with RM HNSCC. The robustness of this method should be further determined in a larger-scale study.

In addition to serving as a novel companion diagnostic, our biomarker exploration provided evidence for the development of more effective therapeutic strategies for non-survivors. PEX *HLA-E* mRNA showed the most prominent role among the 6 candidate PEX mRNAs and a positive correlation with the HLA-E protein expression in the corresponding tumor. HLA-E, which is a nonclassical HLA class I molecule, is overexpressed in cancer cells and strongly inhibits the activity of NK cells and CD8^+^ CTL through the interaction with NKG2A surface receptor, forming the HLA-E/NKG2A immune checkpoint (22, 23, 27). It was clearly demonstrated that HNSCC was associated with the highest infiltration of NK and CD8^+^ CTL across the solid tumors (25) and that these immune cells simultaneously expressed NKG2A and PD-L1 protein (23). This finding indicates the frequent formation of dual immune checkpoints (PD-L1/PD1 and HLA-E/NKG2A) in HNSCC, at least in part, accounting for the limited effects of nivolumab. Moreover, in the phase II UPSTREM study, the anti-NKG2A antibody, monalizumab failed to show clinical efficacy for RM HNSSC progressing after platinum therapy (28). In the phase III INTERLINK 1 study, which targeted RM HNSSC patients previously treated with platinum and PD(L)-1inhibitor, monalizumab in combination with cetuximab (an anti-EGFR antibody) also failed to show clinical efficacy (https://yhoo.it/3OPZbGx). It is likely that the effect of targeting one immune checkpoint is canceled by another immune checkpoint. Thus, our strategy to simultaneously target PD-1/PD-L1 and HLA-E/NKG2A immune checkpoints for biomarker-selected patients appears to be more precise and promising (Fig.5D). The safety and efficacy of the combinational administration of durvalumab (an anti-PDL-1 antibody) and monalizumab were confirmed in the Phase II lung cancer study (29). Thus, a prospective clinical study to test our strategy is encouraged.

Unfavorable markers including *ACTB, MPIG6B, TNFRSF13C*, and the NLR, and favorable markers including *RABL2B,* and *ZNF480*, were also in our prognostic biomarker combination. The levels of PEX *ACTB* mRNA are expected to reflect the total amounts of PEX, as mentioned above, being related to proliferative activity and rapid tumor growth (10). The oncogenic and immunogenic functions of MPIG6B are poorly understood, but a recent study identified that this molecule is essential for the induction of megakaryocytes, which are responsible for myelofibrosis (30). TNFRSF13C is expressed in HNSCC tumor-infiltrating lymphocytes (31), and has been identified as an inducer of regulatory T cells in melanoma (32). The correlation of the NLR with ICI response has been investigated in several reports, including some in HNSCC (18, 19). Overall, the reported predictive value of the pretreatment NLR alone is not sufficient, as confirmed in our study, but its utility in combination with other factors was confirmed. RABL2B is a small RAB GTPase. Interestingly, several members of this family of proteins (e.g., RAB27) are known to regulate exosome biogenesis and to promote melanoma progression (33, 34). However, the physiological and pathological functions of RABL2B remain unclear. The zinc finger protein, ZNF480, is reported to be a core transcription factor required for embryonic stem cell differentiation (35), but its oncogenic function is poorly understood. In summary, the precise roles that make these 6 PEX mRNAs good prognostic biomarkers of nivolumab should be investigated in future studies. However, given the reported and predicted functions of each gene, these molecules likely have functions in the oncogenesis and the immune system, when expressed in PEX mRNA-producing cells (e.g., cancer cells) and recipient cells (e.g., immune cells).

In conclusion, we identified a novel reliable prognostic biomarker for nivolumab that can be assessed with a single pretreatment blood test in this pilot study. Further validation in a larger-scale study is encouraged. A prospective study that examines the efficacy of simultaneous administration of nivolumab and monalizumab in the selected patients by the biomarker should also be performed.

## Methods

### Study design

The BIONEXT study is composed of the following two parts (Fig.1).

Part 1: This part included patients with RM HNSCC patients who were treated with nivolumab. Inclusion criteria were age ≧ 20 years; history of platinum agent administration; pathologically confirmed SCC of the nasal cavity, paranasal sinus, nasopharynx, oropharynx, oral cavity, hypopharynx, or larynx that was recurrent or metastatic and not curable by local therapy; an Eastern Cooperative Oncology Group (ECOG) performance status score of 0 or 1; and at least one tumor lesion measurable per Response Evaluation Criteria in Solid Tumors (RECIST) version 1.1 demonstrated by computed tomography imaging within 28 days of registration. The exclusion criteria were history of ICI therapy or any kind of immunotherapy; and active synchronous or metachronous (within 5 years) cancers except for the carcinoma *in situ* (CIS) and early esophageal cancer curable by endoscopic resection. Enrolled patients were treated with nivolumab (240 mg every 2 weeks or 480 mg every 4 weeks), and their responses were evaluated every 8 weeks until progressive disease (PD) was detected. Clinical data were collected through the Viedec4 electronic data capture system constructed and maintained by the Clinical Research Support Center (CReS) Kyushu. The endpoint of this study was the identification of a PEX mRNA signature that could segregate non-survivors from long-term (> 2-year) survivors. Pretreatment plasma samples (5 mL), collected from peripheral blood, were preserved at −80℃ until assays. Selected pilot samples were subjected to comprehensive RNA-seq analysis for the discovery of candidate PEX mRNA markers, and then the prognosis predictive performance of these markers was validated by RT-qPCR assays in the entire cohort. We initially developed a BOR predicting biomarker employing receiver operating characteristic (ROC) curve analyses and then confirmed its power for prognostic prediction by calculating cumulative survival rates and hazard ratios (HRs) between the marker-selected (i.e., high vs low) cohorts. All assays were conducted in compliance with the minimal information for studies of extracellular vesicle 2018 protocol(36) in the laboratory of Showa Denko Materials America under strict quality and quantity control considering future practical use as a companion diagnostic.

Part 2: This part was designed to confirm that the specific PEX mRNA signature could indeed reflect the TIME of the tumors in the same patient and moreover to elucidate the mechanism of action canceling the effects of nivolumab in non-survivors. Paired tumor and plasma samples were collected from 20 treatment-naïve patients who underwent radical surgery at the National Kyushu Cancer Center. Respective frozen and formalin-fixed paraffin-embedded (FFPE) tumor samples were subjected to mRNA-seq and immunohistochemistry (IHC) to score TIME. Concurrently, the PEX mRNA expression profile of the same patient was evaluated by RT-qPCR in reference to the prognostic biomarker genes established in part 1 and patients were stratified into two groups (survivor vs non-survivor signature). Comparing these two cohorts, the TIME score and the biological implication of PEX mRNA signature were investigated.

### Sample collection

Blood samples, taken within 28 days before nivolumab administration, were immediately centrifuged at 1100xg for 10 minutes and 5 ml of plasma samples were dispensed and snap-frozen at −80℃. Sample collection, preservation, and shipment to Showa Denko America were performed by the SRL Inc. (Tokyo, Japan) under restrict quality and temperature management.

### PEX mRNA isolation and sequencing

PEXs were quantitatively isolated from 1 ml of plasma using an ExoComplete isolation tube kit (Showa Denko Materials, Tokyo, Japan), and total RNA was isolated with a MagMax Total Nucleic Acid Isolation Kit (Thermo Fisher, CA) as previously described unless otherwise noted (37). cDNA libraries were prepared using a TruSeq mRNA stranded library kit (Illumina, CA) and sequenced by paired-end read sequencing on a NovaSeq 6000 (Illumina, CA). The obtained raw reads were mapped against the human genome (GRCh38.p13) by hitsat2 and the read counts were obtained by featureCount on a Linux workstation. Differential gene expression analysis was performed by edgeR.

### PEX mRNA RT-qPCR assay

PEX mRNA isolation was conducted as described above. cDNA was synthesized with qScript XLT cDNA SuperMix (Quantabio, MA, USA) following the manufacturer’s protocol. qPCR was performed with SsoAdvanced Universal SYBR Green Supermix (Bio-Rad, CA, USA) in a ViiA 7 Real-Time PCR System (Thermo Fisher Scientific, CA, USA) with the following protocol: 95℃ for 10 min, followed by 40 cycles of 95℃ for 30 s and 65℃ for 1 min and a melting curve analysis. The primer sequences are shown in Supplementary Table S1. Threshold cycle (Ct) values of the marker candidates were normalized to that of the reference gene (*GAPDH*) using the delta Ct method.

### IHC

Human leukocyte antigen E (HLA-E) and programmed death ligand 1 (PD-L1) protein expression levels in the FFPE tumor samples were analyzed using a Ventana Benchmark Ultra slide processor using antibodies against HLA-E (MEM-E/02; Sant Cruz Biotechnology, Inc.) and PD-L1(22C3; PharmDx). The CPS was calculated according to the standard method (5). HLA-E tumor expression was interpreted as strong when more than half of tumor cells was positive, whereas as low when less than half of cells were positive.

### RNA-seq of primary tumor tissues and scoring of the TIME

RNA extracted from the 17 primary tumor tissues was sequenced on a DNBSEQ-G400 sequencer at Beijing Genomics Institutions (Shenzhen, China). The sequenced reads were aligned to the human reference GRCh38 genome by STAR v2.7.9a with Gencode v38 annotations using the supercomputing system SHIROKANE (University of Tokyo). Transcript-per-million (TPM)-normalized read count tables were generated by RSEM. Downstream analyses were conducted using R v4.1.1. (The R Foundation for Statistical Computing). The IFN-γ-signature (the original 6 genes and an expanded 18 genes signature) and the proportions of immune cells in primary tumor tissues were estimated according to the methods in previous reports (6, 7) and CIBERSORTx (https://cibersortx.stanford.edu/) (24). The 17 cases were divided into two groups according to *HLA-E* expression levels based on the median value. The difference in the IFN-γ-signature and the proportions of immune cells between the *HLA-E* high and low groups were examined by Mann-Whitney U tests. The correlations of the detected marker genes between tissue and PEX-mRNA were examined by Pearson correlation tests.

### Statistics

Data analysis was performed using R version 4.1.1 unless otherwise noted. Statistical significance was determined by a *p-*value of < 5% derived from ANOVA or Welch’s t test. The performance of the marker candidates was evaluated by the AUC of ROC analysis by R package pROC. The optimum threshold was obtained based on the point of the ROC curve nearest to the top-left corner and used to calculate sensitivity, specificity, positive predictive value (ppv), and negative predictive value (npv) to characterize the performance of marker candidates. Sparse logistic regression was also employed to further validate the predictive values of the biomarkers (20). Survival endpoints used to analyze the candidate biomarkers were visualized using Kaplan-Meier analysis. The log-rank test was applied to test the differences among survival curves. Cox proportional hazards regression models were used to calculate the HR.

### Study approval

This study was approved by the Institutional Review Board of the National Kyushu Cancer Center (2019–024), and written informed consent was obtained from all patients before enrolment. This study is registered to the UMIN Clinical Trial Registry: UMIN000037029.

### Data availability

All data needed to evaluate the conclusions in the paper are present in the paper and/or the Supplementary Materials. Additional data related to this paper may be requested from the authors.

### AUTHOR CONTRIBUTIONS

Conceptualization; KS, ST, MM; data curation; MM, KS, MS, KY, YU, TN, HU, OT, HU, TS, SK, KT, AW, IO, NM, SI, MT, YA, NH, DS, HO, TF; methodology: KS, ST, TN, TM, MM; investigation: KS, TN, TH,TM; MM Visualization; KS, ST, TN, MM; Funding acquisition; MM; Supervision; ST, MM; Writing; KS, ST, TM, MM

## Data Availability

All data produced in the present study are available upon reasonable request to the authors

## ACKNOWLEDGEMENTS

We thank Shoji Tokunaga (Medical Information Center, Kyushu University) for his advice on the study design (e.g., sample size) and statistical analyses and CreS Kyushu for intensive support.

---

**Supplementary Table S1.** Patient and tumor characteristics.

| Characteristics | *N* = 104 |
| --- | --- |
| Median age (range) , -years | 67.5 (35-82) |
| Sex -no.(%) |  |
| male | 92(88) |
| female | 12(12) |
| ECOG-performance status, -no.(%) |  |
| 0 | 68(65) |
| 1 | 31(30) |
| 2 | 5(5) |
| Extent of disease, -no.(%) |  |
| Locoregionally recurrence only | 60(58) |
| Distant metastasis | 44(42) |
| Primary cancer site, -no.(%) |  |
| Nasal and paranasal cavity | 9(9) |
| Oral cavity | 31(30) |
| Nasopharynx | 7(7) |
| Oropharynx (p16+) | 5(5) |
| Oropharynx (p16-) | 8(8) |
| Hypopharynx | 31(30) |
| Larynx (supraglottic) | 13(13) |
| Platinum agents previously administered, -no.(%) |  |
| CDDP | 86(83) |
| CBDCA | 21(20) |
| CDGP | 4(4) |
| No. of previous lines of systemic cancer therapy, -no.(%) |  |
| 1 | 73(70) |
| 2 | 27(26) |
| 3 | 4(4) |

CDDP, cisplatin; CBDCA, carboplatin; CDGP, nedaplatin

**Supplementary Table S2.** Candidate best overall response predicting biomarkers.

| Comparison | Gene | logFC | logCPM | F | P-value | FDR | AUC |
| --- | --- | --- | --- | --- | --- | --- | --- |
| PR vs. PD | TAF4B | -2.658 | 2.8695 | 13.179 | 0.001 | 0.992 | 0.9545 |
|  | TESK2 | -1.564 | 2.827 | 5.0272 | 0.032 | 0.992 | 0.9545 |
|  | MFSD8 | -1.729 | 3.0082 | 6.7906 | 0.0138 | 0.992 | 0.9545 |
|  | ZNF480 | -1.755 | 3.2126 | 9.8931 | 0.0036 | 0.992 | 0.9394 |
|  | FAM76A | -2.642 | 2.6952 | 11.335 | 0.002 | 0.992 | 0.9394 |
|  | TGIF1 | -1.718 | 3.2614 | 7.0577 | 0.0122 | 0.992 | 0.9318 |
|  | TNFRSF13C | -2.169 | 2.4797 | 6.4285 | 0.0163 | 0.992 | 0.9242 |
|  | LOC283788 | -2.761 | 2.8714 | 11.072 | 0.0022 | 0.992 | 0.9242 |
|  | SLC25A13 | -2.421 | 2.9492 | 9.1698 | 0.0048 | 0.992 | 0.9091 |
|  | HLA-DQA1 | 0.9238 | 3.5784 | 1.8142 | 0.1874 | 0.992 | 0.9015 |
|  | COL10A1 | -2.917 | 3.1403 | 10.181 | 0.0032 | 0.992 | 0.9015 |
|  | RPL23AP7 | 2.1722 | 3.7521 | 12.158 | 0.0014 | 0.992 | 0.8636 |
|  | CD3D | -1.244 | 2.9224 | 2.6487 | 0.1134 | 0.992 | 0.8485 |
|  | TCF7 | -1.224 | 4.5009 | 5.5421 | 0.0248 | 0.992 | 0.8409 |
|  | HLA-E | 3.5849 | 5.1057 | 9.1891 | 0.0048 | 0.992 | 0.8333 |
|  | HLA-DRA | -0.999 | 3.9583 | 2.3496 | 0.1351 | 0.992 | 0.7879 |
| PR vs. SD/PD | MSH2 | 0.9593 | 3.3172 | 4.0467 | 0.0526 | 0.9982 | 0.7424 |
| PR/SD vs. PD | RABL2B | -1.668 | 3.1032 | 9.4917 | 0.0042 | 0.9401 | 0.7424 |
|  | CTSW | 1.7366 | 5.465 | 15.124 | 0.0005 | 0.9401 | 0.7348 |
|  | MPIG6B | 1.9582 | 3.5563 | 9.3287 | 0.0045 | 0.9401 | 0.7121 |

PR, partial response; PD, progressive disease; SD, stable disease; FC, fold change; CPM, count per million; F, F-statistic values of quasi-likelihood F-test; FDR, false discovery rate, AUC, area under the ROC curve

**Supplementary Table S3.** Candidate response-predicting biomarkers (assessed in responder vs. non-responder groups)

| Parameters | AUC | threshold | sensitivity | specificity | ppv | npv |
| --- | --- | --- | --- | --- | --- | --- |
| HLA-E | 0.729 (0.605-0.853) | -0.282 | 0.765 | 0.632 | 0.903 | 0.375 |
| ACTB | 0.673 (0.547-0.800) | 4.123 | 0.659 | 0.632 | 0.889 | 0.293 |
| MPIG6B | 0.672 (0.548-0.797) | -0.92 | 0.612 | 0.684 | 0.897 | 0.283 |
| RABL2B | 0.614 (0.452-0.777) | -7.296 | 0.624 | 0.632 | 0.883 | 0.273 |
| TNFRSF13C | 0.614 (0.480-0.748) | -9.185 | 0.624 | 0.579 | 0.869 | 0.256 |
| ZNF480 | 0.593 (0.451-0.734) | -8.132 | 0.753 | 0.526 | 0.877 | 0.323 |
| NLR | 0.591 (0.464-0.719) | 4.862 | 0.541 | 0.632 | 0.868 | 0.235 |

AUC, area under the ROC curve; ppv, positive predictive value; npv, negative predictive value

**Supplementary Table S4.**
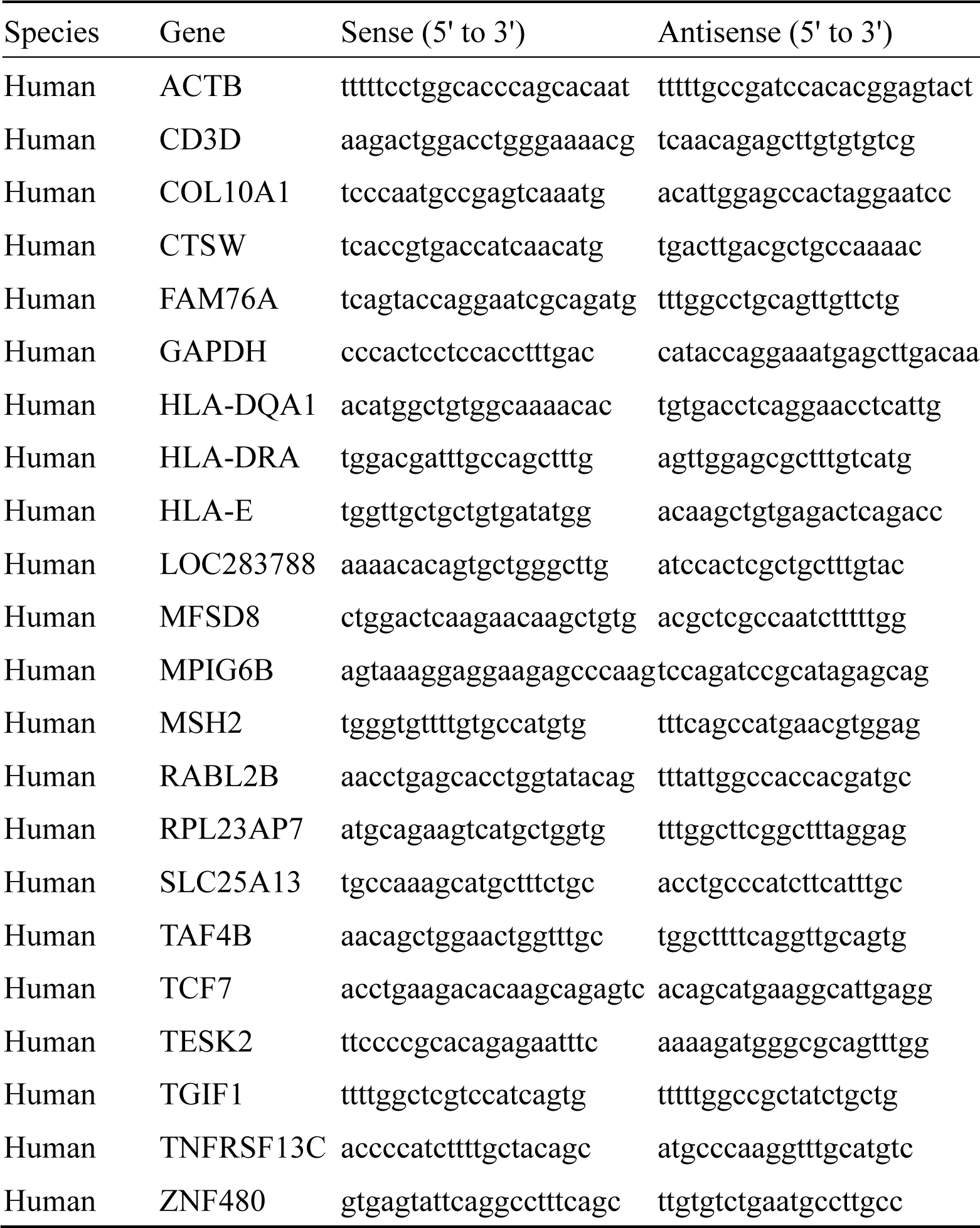
Primer sequences.

